# Use of WATCH antibiotics prior to presentation to the hospital in rural Burkina Faso

**DOI:** 10.1101/2021.02.24.21252387

**Authors:** Daniel Valia, Brecht Ingelbeen, Bérenger Kaboré, Ibrahima Karama, Marjan Peeters, Palpouguini Lompo, Erika Vlieghe, Annelies Post, Janneke Cox, Quirijn de Mast, Annie Robert, Marianne A.B. van der Sande, Hector Rodriguez Villalobos, Andre van der Ven, Halidou Tinto, Jan Jacobs

## Abstract

**Background:** In low- and middle-income countries (LMIC), the prevalence of antimicrobial resistance (AMR) is increasing. WHO recommends monitoring antibiotic use, in particular Watch antibiotics, clinical important but at risk of becoming ineffective due to increasing AMR. We investigated antibiotic use at primary care or community-level in rural Burkina Faso.

**Methods:** During 2016-2017, patients aged >3 months admitted with severe acute fever to the rural hospital of Nanoro Health District, Burkina Faso, reported antibiotic use in the two weeks prior to consultation or hospitalization, which we analysed using the WHO Access, Watch, Reserve (AWaRe) classification. Most Watch antibiotics, e.g. ceftriaxone, are not recommended at primary health center level, as is also the case for ciprofloxacin in children.

**Results:** Of 920 participants (63.0% ≤14 years), pre-admission antibiotic use was reported by 363 (39.5%) of whom 58 (16.0%) reported more than one antibiotic. Use was more frequent among health center referrals (231, 54.0%) than among self-referred patients (131, 26.7%, p<0.001). Of 424 antibiotics, 261 (61.6%) were Access and 159 (37.5%) Watch antibiotics. Watch antibiotics use was more frequent among >14 year olds (72, 51.1%) than 0-14 year olds (87, 30.7%) and among referrals (41, 28.1%) compared to self-referred patients (117, 42.2%). Most frequently used Watch antibiotics were ceftriaxone (114, 26.9%) and ciprofloxacin (32, 7.5%). Among antibiotics reported by referral patients, ceftriaxone and ciprofloxacin were respectively recorded 100 (36.1%) and 12 times (4.3%).

**Conclusion:** The frequent use of Watch group antibiotics prior to presentation to the hospital in rural Burkina Faso highlights the need to address primary care, over-the-counter and informal community-level antibiotic use as part of antibiotic stewardship in LMIC, facilitating referral, access to qualified prescribers, or improving diagnostic tools in health centers.

## Introduction

The emergence of antimicrobial resistance (AMR) is driven by appropriate and inappropriate use of antimicrobials. Increasing microorganisms’ exposure to antibiotics results in selection pressure, while suboptimal dosage allows selective survival of resistant microorganisms [1]. Globally, the burden of AMR infections has been estimated to increase from 700,000 annual deaths in 2014 to 10 million in 2050 [2]. The World Health Organization’s Global Action Plan on AMR from 2015 urges to generate knowledge on AMR and antibiotic use, and to optimize the use of antibiotics [3]. A global surveillance network for monitoring AMR and antibiotic use, the Global Antimicrobial Resistance Surveillance System (GLASS), has since been set up, reporting country-wide AMR prevalence of key human pathogens and antibiotic use [4]. To monitor the antibiotic use, WHO proposed a classification of antibiotics for human use in three groups, “Access”, “Watch” and “Reserve”, according to their clinical importance, specific recommendations for their appropriate use, and resistance potential [5]. Antibiotic sales data during 2000-2015 showed that the human use of so-called Watch antibiotics, critically important antibiotics particularly at risk of AMR emergence, was declining in high-income countries, while importantly growing in middle income countries for which data were available [6,7].

In sub-Saharan Africa, self-medication with over-the-counter antibiotics from private pharmacies or informal drug stores, without prior prescription by a qualified health worker, is frequent and facilitates inappropriate antibiotic use [8–10]. To optimize (inappropriate) antibiotic use, it is crucial to understand the role of antibiotic use in the community, outside the hospital setting. Furthermore, even in local primary health centers, antibiotic prescriptions are not always rational [11,12]. Peripheral health workers’ poor understanding of the risk of AMR, limited diagnostic tools and difficult access to qualified referral healthcare are likely to be fuelling irrational prescribing of antibiotics [13,14]. National level antibiotic consumption estimated from wholesale data, including Burkina Faso, are limited to the official healthcare sector and aggregate in- and outpatient use [13]. To our knowledge, no studies have investigated community-level antibiotic use. The purpose of this study is to describe community-level and primary care antibiotic use in febrile patients prior to presentation to the hospital in a rural district of Burkina Faso.

## Methods

### Study population

From March 23, 2016 to June 30, 2017, the PaluBac study recruited patients aged >3 months presenting to the Nanoro district hospital with acute fever (tympanic temperature ≥ 38.0 ° C) or history of fever in the last 48 hours and ≥1 symptom(s) of respiratory distress, generalized weakness (prostration), impaired consciousness, seizures (one or more episodes), clinical jaundice, signs of shock, or with suspected severe malaria, invasive bacterial infection, or severe viral infection. From March 23 to November 8, 2016, only inpatients were recruited; from November 9, 2016 to June 30, 2017 also outpatients were included. PaluBac evaluated the performance of an automated cell counter to quantify malaria parasitaemia [16]. Our embedded cross-sectional study recorded patients’ antibiotic use before presenting to the district hospital.

The study was conducted in the Nanoro Health District in the West-Central region of Burkina Faso, about 90 km from Ouagadougou, the capital city of Burkina Faso. The district has 24 primary health centers and one district hospital, “Centre Médical avec Antenne Chirurgicale” (CMA). Primary health centers are meant to be the patients’ first point of contact with the healthcare system, during outpatient consultations. The only diagnostic tools available at this level of care are malaria rapid diagnostic tests (RDT). There are no medical doctors. The use of ceftriaxone is recommended at the primary health centers level only in case of a meningitis outbreak [17]. More complicated cases are referred from the primary health centers to the district hospital [14]. The district hospital has hospitalization units, medical doctors and a clinical microbiology laboratory. In the health district, the most frequently reported diagnosis is malaria [18], though in children often associated with a community-acquired invasive bacterial co-infection [19,20]. Malaria is endemic with a *high malaria transmission season* from July to November. This period overlaps with the rainy season, which extends from June to October. The *low malaria transmission season* is from December to June.

### Data collection

At study inclusion, study clinicians completed case report forms with medical history, including the use of antibiotics and antimalarials in the two weeks prior to consultation or admission at the district hospital, which we refer to as *pre-admission antibiotic use*, the date of symptom onset, and whether the patient was referred or not from a primary health center. *Referral patients* first attended a primary health center where a nurse decided on the need for higher-level care and referral to the district hospital, based on clinical symptoms. Inpatient care is not possible at a primary health center, the patient can only be observed for a maximum of 48 hours. *Self-referred patients* were those presenting directly at the district hospital without first attending a primary health center.

### Data analysis

Pre-admission antibiotics were classified (i) according to WHO’s Access, Watch, Reserve (AWaRe) classification and (ii) according to products name as stated in the WHO essential medicines list [5]. For the latter, we combined ampicillin and amoxicillin use into one category (“ampi/amoxicillin”), and zoomed in on the use of the most prevalently prescribed antibiotic in the Watch group in particular. Anti-tuberculosis drugs were excluded from analysis. We report frequencies of antibiotics used by product name and by AWaRe group, of the use of more than one antibiotic, and compared by age (0-14 years vs >14 years), by malaria transmission season, by hospitalisation status (in-vs outpatients) and by referral vs self-referred, using chi-square tests. We stratified the frequencies of pre-admission antibiotic use by malaria transmission season. Data was analysed with Stata 14.

### Ethics

The PaluBac study protocol and an amendment to include pre-admission antibiotic use were approved by the Ethics Committee for Health Research of Burkina Faso (ref 1029/15 and 2017-01-001). Informed consent was obtained from all included patients or their caretakers.

## Results

### Characteristics of the study population

Of 1212 screened patients, 920 (75.9%) patients had data on antibiotic use recorded. The median age was 5 years (inter-quartile range, IQR, 1-30). 580 (63.0%) were children aged 0-14 years and 533 (57.9%) were men. A total of 428 (46.5%) patients were referrals from primary health centers; 344 (37.4%) patients were hospitalized upon arrival to the district hospital.

### Pre-admission antibiotic use

Pre-admission antibiotic use was reported by 363 (39.5%) patients, of whom 58 (16.0%) used more than one antibiotic. Pre-admission antibiotic use was reported by 240 (41.4%) of 0-14 year olds and 123 (36.2%) of >14 year olds (p=0.12). Referral patients more frequently reported pre-admission antibiotic use (231, 54.0%) than those self-referred (131, 26.7%, p<0.001). Fewer patients (131, 34.7%) reported antibiotic use during the high malaria transmission season than outside (232, 42.7%, p<0.001). During the first recruitment period (from March 23 to November 8, 2016), 121 patients (35.2%) reported antibiotic use and during the second recruitment period (from November 9, 2016 to June 30, 2017), antibiotic use was reported by 242 patients (42.0%) (p=0.04), but when stratifying by malaria transmission season, we did not find a difference in antibiotic use between both recruitment period (p=0.63).

### AWaRe distribution of antibiotics

Overall, 424 antibiotics were reported: 261 (61.6%) Access, 159 (37.5%) Watch and no Reserve group antibiotics. Also 4 (0.9%) unclassified antibiotics were reported. Watch antibiotics were more frequently reported by >14 year olds (72, 51.1%) than 0-14 year olds (87, 30.7%, p<0.001) and by referrals (117, 42.2%) compared to self-referred patients (41, 28.1%, p=0.004). There was no difference in the proportion of Watch antibiotics between high and low malaria-transmission season (Table 1).

**Table 1:**
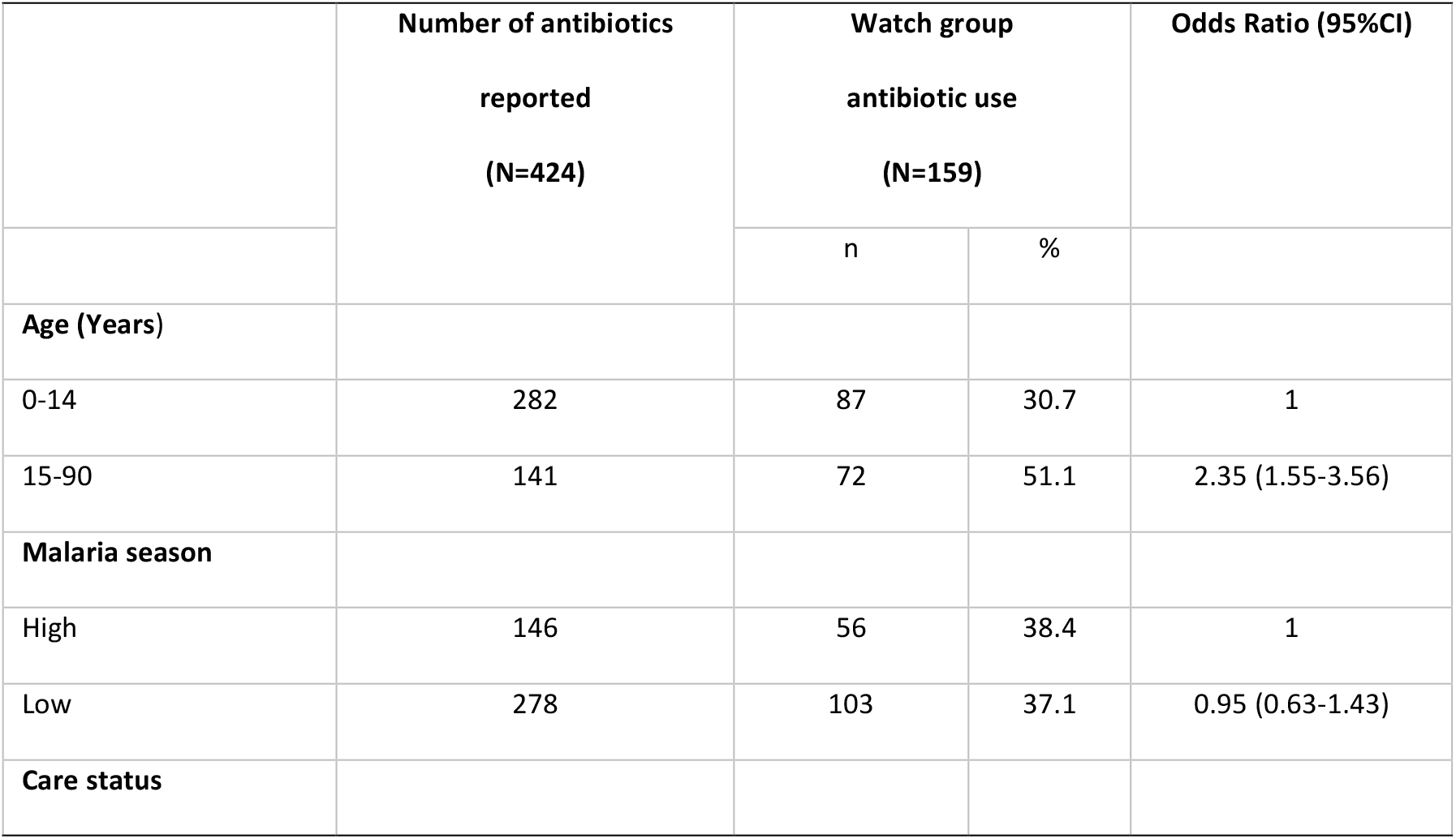
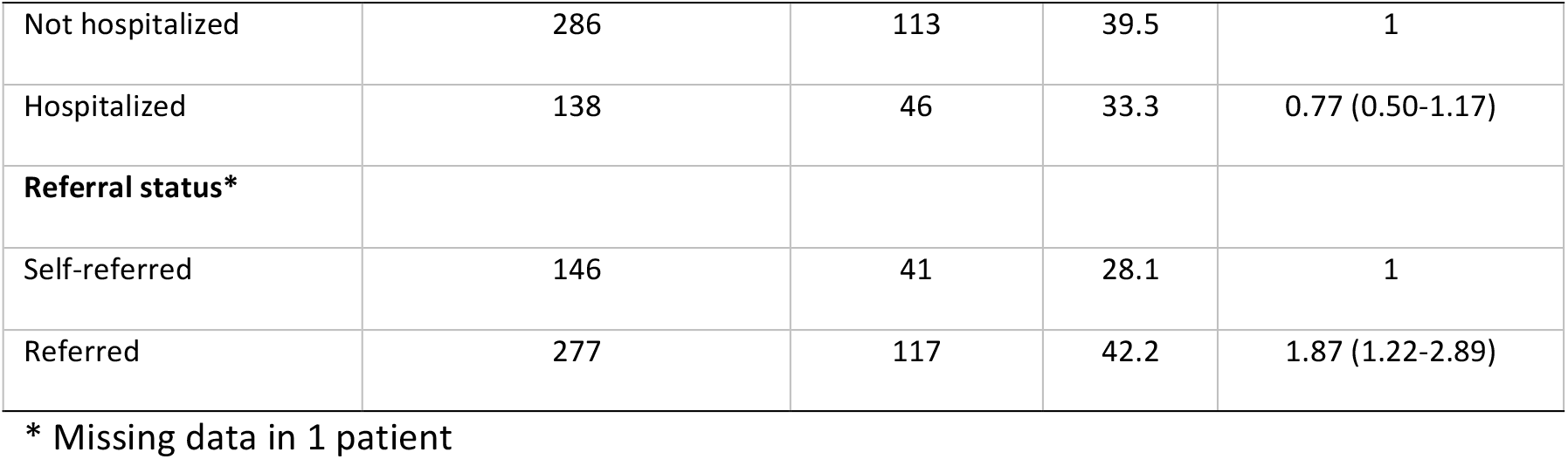
Factors associated with pre-admission Watch group antibiotic use, from March 23, 2016 to June 30, 2017 at the Nanoro district hospital (424 antibiotics used by 363 patients reporting antibiotic use)

|  | Number of antibiotics reported<br>(N=424) | Watch group antibiotic use<br>(N=159) |  | Odds Ratio (95%CI) |
| --- | --- | --- | --- | --- |
|  |  | n | % |  |
| **Age (Years)** |  |  |  |  |
| 0-14 | 282 | 87 | 30.7 | 1 |
| 15-90 | 141 | 72 | 51.1 | 2.35 (1.55-3.56) |
| **Malaria season** |  |  |  |  |
| High | 146 | 56 | 38.4 | 1 |
| Low | 278 | 103 | 37.1 | 0.95 (0.63-1.43) |
| **Care status** |  |  |  |  |
| Not hospitalized | 286 | 113 | 39.5 | 1 |
| Hospitalized | 138 | 46 | 33.3 | 0.77 (0.50-1.17) |
| **Referral status\*** |  |  |  |  |
| Self-referred | 146 | 41 | 28.1 | 1 |
| Referred | 277 | 117 | 42.2 | 1.87 (1.22-2.89) |

\* Missing data in 1 patient

### Antibiotics reported

Ampicillin or amoxicillin use was most reported (access group, 137, 32.3%). The most frequently used Watch antibiotics were ceftriaxone (114, 26.9%) and ciprofloxacin (32, 7.5%). Among antibiotics reported by referral patients, ceftriaxone was recorded 100 times (36.1%) and ciprofloxacin 12 times (4.3%). Among antibiotics reported by self-referred patients, ciprofloxacin was recorded 20 times (13.7%) (Fig 1).

**Fig 1.**
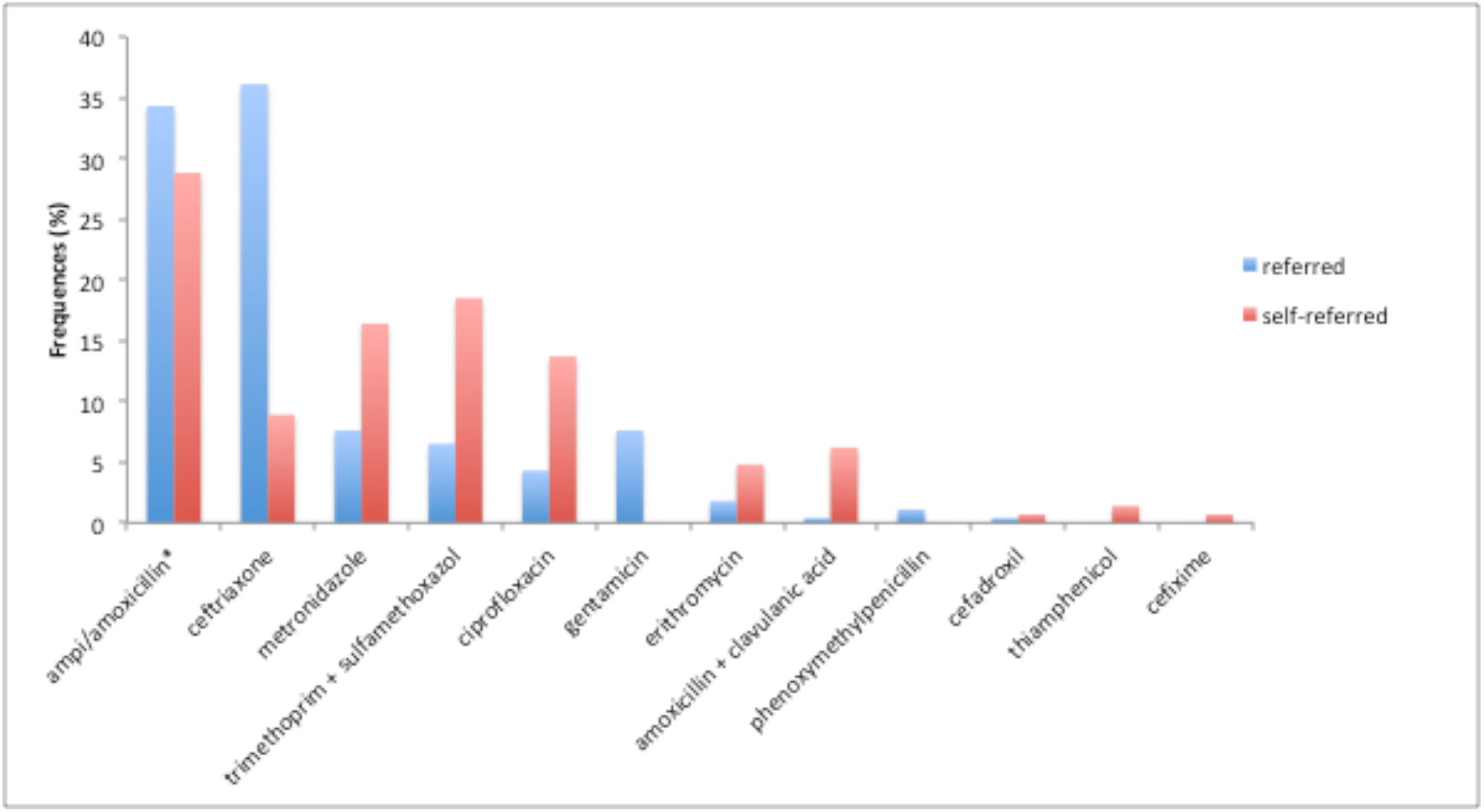
Frequency of antibiotics used prior to admission or consultation for severe febrile illness at Nanoro district hospital, March 2016-June 2017. Frequency of an antibiotic reported among patients referred from primary health centers is expressed as the number of this antibiotic divided by the total number of all antibiotics reported in referred patients (n = 277). Frequency of an antibiotic reported among self-referred patients is expressed as the number of this antibiotic divided by the total number of all antibiotics reported in self-referred patients (n = 146). *Ampi/amoxicillin is use of either ampicillin or amoxicillin Ampicillin, ceftriaxone and gentamicin were intravenously administered. Of the reported antibiotics, 196 (46.2%) were parenteral versus 228 (53.8%) oral. Parenteral antibiotic use was slightly more frequent among 0-14 year olds (48.4%) than >14 year olds (41.8%, p=0.20).

## Discussion

Nearly 40% of patients presenting with severe fever to a referral hospital in rural Burkina, reported preceding antibiotic use; more than half of referrals but also a quarter of self-referred patients. While empirical first- or second-choice (Access) antibiotics were most frequently used, it was surprising that Watch antibiotics were used by 42% of patients referred from a primary health center. This is noteworthy particular since ceftriaxone is not on the Burkina Faso essential medicines list for primary health center level [17]. Moreover, Watch antibiotics accounted for 28% of antibiotics used by self-referred patients, who probably self-medicated with antibiotics obtained without prescription at private pharmacies or from informal drug sellers. The high proportion of ceftriaxone – administered parenteral – among the Watch antibiotics reported by the non-referral patients is striking.

High and increasing use of Watch antibiotics has been observed in other low- and middle-income settings [21–23]. To introduce interventions to optimize antibiotic use, it is important to understand the origin and reasons why Watch antibiotics are used, both at primary health center level and without prescription at community-level. Our study illustrates the need to monitor antibiotic use by official and by informal healthcare providers. Nationwide antibiotic consumption estimated from official sales data, showed that in 2015, 75% antibiotics consumed in Burkina Faso were Access and 24% Watch group, attaining the WHO target of at least 60% Access antibiotics used [24]. This is lower than the high proportion of Watch antibiotics use observed in the present study, which did not include inpatient antibiotic use, where a higher proportion of Watch antibiotics can be expected. In comparison, the frequency of pre-admission antibiotic use by febrile patients admitted to the Nanoro district hospital in 2012-2013 was lower (28.2%), and the proportion of Access antibiotics used was higher (45.9% amoxicillin or ampicillin vs 32.3% now and 35.1% TMP-SMX vs 10.6% now)[25]. In another study on paediatric (6-59 months) antibiotic use at primary health center level in the rural Nouna district of Burkina Faso in 2017, use of Watch group antibiotics was less frequent (16.4%) and mainly consisted of erythromycin, an antibiotic authorized at this level [26].

Despite ampicillin being the treatment of choice for patients with severe infectious diseases at primary health center level pending referral, it is increasingly ineffective against *Enterobacterales*. Indeed, most (90.5%) of non-Typhi *Salmonella* and 87.5% of *E. coli* were resistant to ampicillin in Nanoro during 2012-2013 [25]. This could explain (systematic) use of ceftriaxone when bloodstream infection is suspected in the absence of a microbiological testing. Rapid referral to the district hospital, where laboratory testing should be available as recommended in the WHO model list of essential in vitro diagnostics [27], is preferable. Because cephalosporin use has been associated to the emergence of beta-lactamase-secreting pathogens, particular caution must be taken to optimize their use [27]. Integrating point-of-care CRP or procalcitonin tests at primary health center level, to direct bacterial and non-bacterial fever, could be explored.

Some caution interpreting these findings is needed. First, data about antibiotic use were available for slightly more than three-quarter of patients. Further, we included only patients presenting with severe acute fever, not representative of any illness episode in the community. Nevertheless, no meningitis outbreak was reported during the study period, so ceftriaxone use was not recommended at the primary health center level [17]. Antibiotic use was collected in a survey, recalling use during two weeks before admission to the district hospital, potentially underestimating actual use. Whenever possible, reported antibiotic use was verified from referral forms (if referral), patient medical files (healthcare booklet), and antibiotic packaging or blisters. The inclusion of outpatients during the second half of the study period might have resulted in some changes in the study population and pre-admission antibiotic use. However, we found no difference in the prevalence of antibiotic use, when comparing the two populations from both recruitment periods (p=0.63).

## Conclusion

The elevated use of Watch antibiotics in primary health care centers and at community-level in rural Burkina Faso is concerning in a context of ineffective first-line antibiotics and increasing detection of beta-lactamase-secreting bacteria. It points to the need to set up antibiotic use monitoring and antibiotic stewardship in rural communities in LMIC, as over-the-counter or informal community-level antibiotic use can be very frequent in communities with limited access to hospital care. As no alternatives for the currently available Watch antibiotics are available nor affordable in most LMIC settings, it is imperative to act to avoid an “epidemic of resistance” in years to come.

## Data Availability

Data used for the different analyses in the manuscript are available and may be share if needed

## Acknowledgments

We acknowledge all the Palubac study team and all study participants for their willingness to participate in this study. We are also thankful to the Palubac study staff, mainly the clinicians and nurses who participated in data collection. We thank all the Clinical Research Unit of Nanoro staff as well as the Nanoro district hospital staff for being part in some ways of the success of this study.

## Authors’ contributions

**Conceptualization:** DV, BI, MvdS,JJ

**Data curation:** DV

**Formal analysis:** DV,

**Investigation:** BK, IK, MP, PL, AP, JC

**Methodology:** DV, BI, MvdS, JJ

**Software:** DV

**Supervision:** HT, MABvS, JJ

**Validation:** HT, MABvdS, JJ

**Writing – Original Draft Preparation:** DV

**Writing – Review & Editing:** BI, BK, MP, PL, EV, AP, JC, QdM, AR, MABvdS, HRV, AvdV, HT, JJ

